# Competing event regression on the relative subdistribution and cumulative-incidence scales

**DOI:** 10.64898/2026.08.13.26360204

**Authors:** Loren K. Mell, Morten Overgaard

## Abstract

In competing risks settings, covariate effects and group comparisons are usually assessed one event at a time, through log-rank or Cox tests on the cause-specific hazards, or Gray’s test or Fine–Gray regression on a cumulative incidence function (CIF). This can obscure a clinically important quantity: the ratio between the event of interest and the competing event, since groups may differ little on the individual events yet differ sharply in their ratio. The generalized competing event (GCE) framework makes this ratio the object of inference; on the cause-specific scale the hazard ratio *ω*^+^(*t*) = *λ*_1_(*t*)*/λ*_2_(*t*) is estimated efficiently from a single stacked (Lunn–McNeil) model. We extend the framework to two scales that describe realized incidence. The *subdistribution hazard ratio* 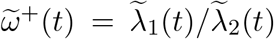 is estimated by a stacked, risk-set-weighted extension of the Lunn–McNeil construction; the *cumulative-incidence ratio ρ*(*t*) = *F*_1_(*t*)*/F*_2_(*t*) by a stacked per-cause pseudo-observation regression of the Aalen–Johansen estimator. We relate the three contrasts: *ρ* equals *ω*^+^ exactly under proportional cause-specific hazards, and equals 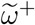 only in the small-time limit under proportional subdistribution hazards, drifting toward 1 thereafter. The orthogonality that makes *ω*^+^ efficient is lost on both cumulative-incidence scales: 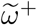 through overlapping weighted risk sets and shared censoring weights, and *ρ* through the shared survivor. Each carries a covariance term that must be handled and bounds efficiency relative to the hazard-scale test. We derive the corresponding variances, study operating characteristics by simulation, illustrate on hypothetical clinical cohorts, and provide an implementation in the gcemod R package.

## 1 Introduction

Competing risks arise when the occurrence of a primary event (or set of events) of interest can be precluded by another event (or set of events). Two regression frameworks dominate practice: the cause-specific hazards model, which treats competing events as censored, and the subdistribution hazards model, which links covariates directly to the cumulative incidence function (CIF)^1,2^. In most applied analyses, covariate effects and group comparisons are examined one event at a time. This is appropriate when interest lies in the individual endpoints, but can be misleading when the clinically relevant contrast concerns the *ratio* between the event of interest and the competing event, rather than each (set of) event(s) individually.

The generalized competing event (GCE) framework makes this ratio the object of inference^3,4^. On the cause-specific hazard scale, it targets *ω*^+^(*t*) = *λ*_1_(*t*)*/λ*_2_(*t*), the ratio of the hazard for the event of interest to that for the competing event, and the related share *ω*(*t*) = *λ*_1_(*t*)*/*(*λ*_1_(*t*) + *λ*_2_(*t*)). These have been used to select patients for treatment intensification in head-and-neck and prostate cancer^5,6^. Cause-specific GCE effects can be estimated efficiently from a single stacked (Lunn–McNeil) model^7^. Because the two cause-specific counting processes have no common jumps, their score processes are orthogonal, and log *ω*^+^ = *β*_1_ *− β*_2_ is a difference of two uncorrelated log-hazard-ratios.

However, clinicians and researchers are often interested in *realized incidence*, i.e. the cumulative probabilities *F*_*k*_(*t*), and the subdistribution hazards that give rise to them, in addition to instantaneous cause-specific rates. It is therefore natural to ask whether the GCE framework can be defined and tested on these scales. We develop two such estimators:

1. the *subdistribution hazard ratio* 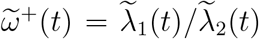, the ratio of the two Fine–Gray subdistribution hazards, estimated by a risk-set-weighted, stacked extension of the Lunn–McNeil construction (Section 3.1); and
2. the *cumulative-incidence ratio ρ*(*t*) = *F*_1_(*t*)*/F*_2_(*t*) (equivalently, the odds that a subject’s realized event by *t* is the event of interest) estimated by a stacked per-cause pseudo-observation regression (Section 3.3).

Existing cumulative-incidence quantities include the Pepe–Mori conditional probability *F*_1_*/*(1*− F*_2_)^8^ and the vertical-modeling relative cause probabilities *P* (cause = *k* | *T* = *t*) ^9^. Both differ from the contrasts we study, as we discuss in Section 7. A single theme organizes the methodology: the orthogonality that makes the cause-specific *ω*^+^ efficient is lost on both cumulative-incidence scales, so each new estimator carries a covariance between the two causes that must be handled in the variance, which bounds its efficiency relative to the hazard-scale test. A single design likewise unifies the estimators: on all three scales the contrast is read off a stacked fit – an event-type indicator, nuisance main effects, and interaction coefficients that are the contrast – with a subject-level variance. In this paper, we make this precise for each estimator, relate the three contrasts, and quantify the resulting trade-off between interpretability and efficiency.

## 2 Three scales for the generalized competing event contrast

Consider two mutually exclusive event types, the event of interest (*k* = 1) and the competing event (*k* = 2), with right censoring. Let *λ*_*k*_(*t* | *z*) be the cause-specific hazards, 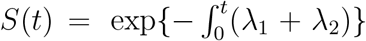 the all-cause survivor, and 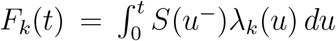 the CIFs. The subdistribution hazard for cause *k* is 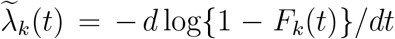, so that 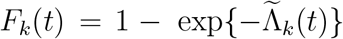 with 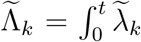. The GCE contrast admits a definition on each of the three scales:

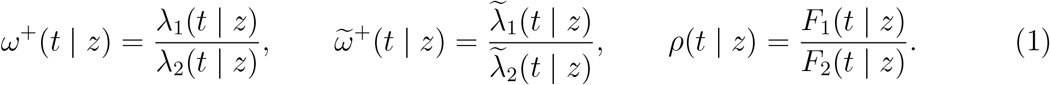

The cause-specific *ω*^+^ contrasts instantaneous rates among those still at risk; the subdistribution 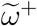 contrasts the hazards that drive the two CIF trajectories; and *ρ* contrasts the realized cumulative probabilities themselves. All three are functionals of the observable competing-risks data.

### 2.1 The cause-specific scale and treatment selection

The cause-specific scale carries a decomposition that motivates the framework and is not reproduced on the other two scales. Suppose an intervention acts multiplicatively on each cause-specific hazard, *λ*_*k*_ *↦→ θ*_*k*_*λ*_*k*_, with *θ*_*k*_ the cause-specific hazard ratio for cause *k*. The all-event hazard *λ*_1_ + *λ*_2_ then has hazard ratio

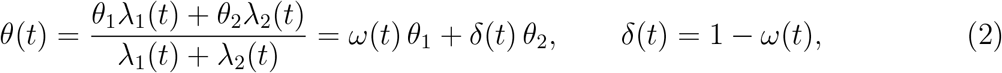

the hazard-share-weighted average of the two cause-specific effects^10,11^, in which *δ*(*t*) is labeled the *deadweight* factor, i.e. the share of the event hazard borne by competing events, which a therapy directed at the event of interest cannot reduce. When the two events partition all-cause mortality, *θ*(*t*) is the effect on overall survival. Equation (2) is the basis for using *ω* (equivalently *ω*^+^) to guide treatment selection: a therapy directed at the event of interest typically has *θ*_1_ *<* 1 while a treatment that confers no benefit on the competing event, *θ*_2_ *≥* 1; then *θ*(*t*) = 1 *−ω*(*t*)(1 *−θ*_1_) + *δ*(*t*)(*θ*_2_ *−* 1) is decreasing in *ω*(*t*), so the composite benefit is largest for patients whose risk is dominated by the event of interest^5,6^. Under the a priori bound (*θ*_2_ *≥* 1) the deadweight factor bounds the composite effect from below, regardless of the treatment effect on the event of interest. Two features of (2) bear on what follows. First, *ω* is a genuine effect modifier of the composite treatment effect, not merely a prognostic index. Second, no comparably clean decomposition holds on the subdistribution or cumulative-incidence scales, because *F*_*k*_ is a nonlinear functional of both hazards and the shared survivor. Effects on 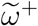 and *ρ* therefore need not track the cause-specific picture, which is why they are worth examining as complementary summaries of the same balance.

### 2.2 The cumulative-incidence ratio

The CIFs are *additive*:

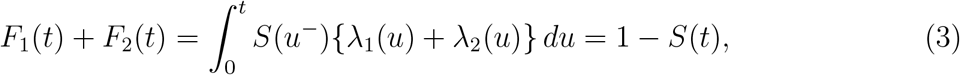

because each is an integral against the same *S*. Additivity makes the share *π*_1_(*t*) = *F*_1_(*t*)*/{F*_1_(*t*)+ *F*_2_(*t*)*}* = *F*_1_(*t*)*/*(1 *−S*(*t*)) a proper probability, i.e. the probability that, among subjects who have experienced any event by *t*, the event was the event of interest, and

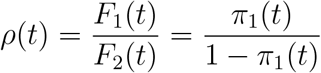

is the corresponding odds, with log *ρ*(*t*) = logit *π*_1_(*t*). Thus *π*_1_(*t*) parallels the instantaneous share *ω*(*u*) and *ρ*(*t*) parallels *ω*^+^(*u*), one scale up in cumulation.

Writing *λ*_1_(*u*) = *ω*^+^(*u*)*λ*_2_(*u*) and substituting into *F*_1_ = ∫ *Sλ*_1_ gives the exact identity

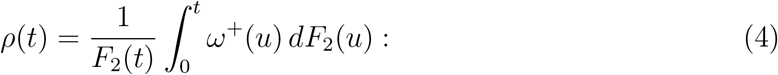

i.e., *ρ*(*t*) is a *dF*_2_-weighted time-average of the instantaneous cause-specific hazard ratio.

### 2.3 Relating the three contrasts

The two hazard-scale contrasts are the instantaneous limits of *ρ* under their respective proportional-hazards assumptions, but they bind to *ρ* differently. The proportionality conditions in this subsection concern the relative hazards under baseline conditions, i.e. whether *ω*^+^(*t*) (respectively 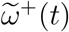) is constant in *t* at fixed covariate values. These are distinct from the modeling assumption of Section 3, under which it is the covariate *contrast* on the relative hazard (the ratio of relative hazards across levels of *z*) that is taken time-constant.

#### Proposition 1

(Cause-specific). *If the baseline cause-specific hazard ratio is constant in time, ω*^+^(*u*) *≡ ω*^+^, *then ρ*(*t*) = *ω*^+^ *for all t, and S cancels*.

This is immediate from (4). Under proportional cause-specific hazards the ratio of the *actual* cumulative incidences equals the cause-specific hazard ratio at every time and does not drift.

#### Proposition 2

(Subdistribution). *If the baseline subdistribution hazard ratio is constant in time* 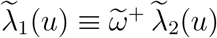, *then* 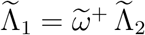 *and*

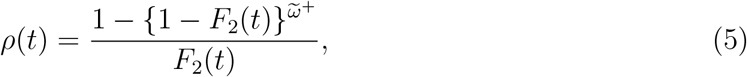

*a function of the accrued competing incidence F*_2_(*t*) *that moves monotonically from* 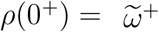 *toward* 1 *as F*_2_(*t*) *increases*.

The proof substitutes 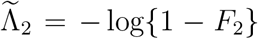 into 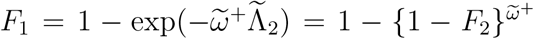. Under proportional subdistribution hazards *ρ* therefore coincides with 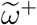 only as *t →* 0 and is then drawn toward the null as incidence accumulates, in contrast to the exact, time-invariant identity *ρ ≡ ω*^+^ under proportional cause-specific hazards. The magnitude of this drift is set by the accrued competing incidence: the more of *F*_2_ realized by time *t*, the closer *ρ*(*t*) is pulled to unity. Thus *ρ* departs markedly from 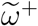 in high-incidence cohorts yet stays close to it when incidence is low (Section 5). The null value 1 is approached only in the limit *F*_2_ *→* 1; whenever the competing event is genuinely improper, *F*_2_(*∞*) = *p*_2_ *<* 1, so *ρ* is drawn only to 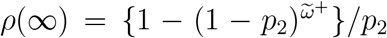 and remains bounded away from unity. Exact convergence to the null coincides with the degenerate limit in which the competing event becomes certain and no genuine competition remains. Whenever both events retain positive eventual incidence, *ρ* is bound more tightly to the cause-specific *ω*^+^ than to the subdistribution 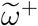 and is held away from the null by the incidence still unrealized. Thus *ω*^+^, 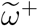, and *ρ* are genuinely distinct summaries of the same balance, and reporting more than one is informative rather than redundant.

### 2.4 Proportional relative hazards under standard assumptions

The estimator of Section 3 assumes a time-constant covariate contrast, equivalently a time-constant relative-hazard ratio RHR(*t*) = *ω*^+^(*t* | 1)*/ω*^+^(*t* | 0). This is not automatic, but under the proportionality ordinarily assumed it reduces to a familiar condition. If the composite and primary-event hazards are proportional across levels of a covariate or treatment, with hazard ratios *θ* and *φ* (*θ≠ φ*), then subtraction gives HR_2_(*t*) = *θ* + (*θ − φ*) *ω*^+^(*t* | 0) for the competing event and RHR(*t*) = *φ/*HR_2_(*t*), whence

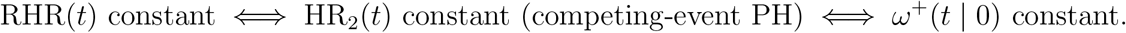

Given proportional composite and primary hazards, then, proportional relative hazards, proportional competing-event hazards, and a time-constant baseline relative hazard are one and the same condition.

### 2.5 Interpretation and non-collapsibility

Two distinct properties govern how a covariate effect on *ρ*(*t*) (and, in parallel, on 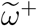) should be read. First, *ρ*(*t*) is *marginal over time*: by (4) the survivor *S* enters through the weights, so a covariate that changes overall event timing can move *ρ*(*t*) even when it leaves the instantaneous balance unchanged. This is a feature of the estimand’s cumulative construction (Propositions 1–2), not a statistical artifact, and it is why *ρ*(*t*) must be reported at a stated time *t*.

Second, and separately, the effect measures are *non-collapsible*. Because log *ρ*(*t*) = logit *π*_1_(*t*), a coefficient on log *ρ* is a conditional log-odds-ratio for the event type among realized events; the subdistribution and cause-specific coefficients are log-hazard-ratios. As for the odds ratio and hazard ratio generally, the conditional effect does not in general equal the covariate-averaged marginal effect, and coefficients are not directly comparable across models with different adjustment sets. This is distinguished from bias, since the estimators remain consistent for the conditional estimands they target^12^. The property is shared by all three GCE contrasts, so it introduces no interpretive issues beyond those acceptable for Cox and Fine–Gray regression. Under the null of no covariate effect, the conditional and marginal measures coincide, so the level and validity of the tests are unaffected. Non-collapsibility concerns the interpretation of effect magnitude, not the inference on which the GCE frame-work rests. Accordingly, each 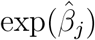 should be reported as a conditional association given the other covariates. When a collapsible summary is desired, a difference of CIFs (or of shares *π*_1_) is collapsible, and a standardized (g-computed) marginal *ρ*(*t*) contrast is available within the same pseudo-observation framework, formed by predicting 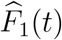 and 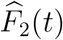 under fixed covariate values and averaging over the empirical covariate distribution.

## 3 Estimation

### 3.1 Stacked Lunn–McNeil estimation of the subdistribution ratio

The Fine–Gray model^1^ specifies proportional subdistribution hazards 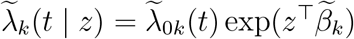, so that the GCE contrast on this scale is

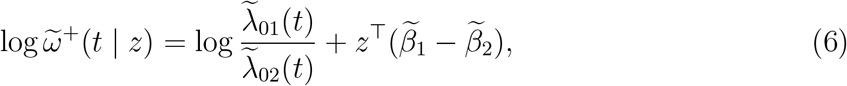

and the covariate effect on 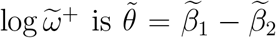. A separate Fine–Gray fit for each cause yields 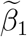 and 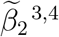, but a single stacked model gives the contrast and an omnibus test directly, extending the cause-specific Lunn–McNeil construction^7^ to the subdistribution scale.

For each cause *k*, form the risk-set-weighted data in which every subject remains in the cause-*k* subdistribution risk set until administrative follow-up, weighted by *w*_*ki*_(*t*) = *Ĝ* (*t*)*/ Ĝ{*min(*T*_*i*_, *t*)*}* with *Ĝ* the Kaplan–Meier estimate of the censoring survival^13^. Stack the two weighted data sets, label them by an event-type indicator, and fit one weighted Cox model stratified by event type with the covariates *z* and their interactions with the indicator. Writing 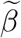 for the main effects (cause 2) and 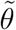 for the interactions, the interaction coefficients estimate 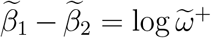 per covariate, and a multivariate Wald test of 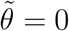 is the omnibus GCE test on the subdistribution scale. This is the direct analogue of the cause-specific stacked estimator, with risk-set weighting in place of censoring-at-competing-event. Earlier GCE analyses estimated this subdistribution contrast covariate-by-covariate from separate Fine–Gray fits^3,4^, obtaining contrast confidence intervals by bootstrap resampling^4^; the stacked fit yields the contrast and, as the next section shows, an analytic variance directly.

### 3.2 Variance of the stacked subdistribution estimator

On the cause-specific scale the stacked estimator has a clean variance: the two counting processes *N*_1_, *N*_2_ have no common jumps, so the score martingales ∫ (*dN*_*k*_ *−Y λ*_*k*_ *du*) are orthogonal, the cause-specific estimators are asymptotically uncorrelated, and 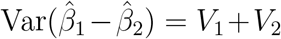 with no covariance term. However, this orthogonality does not pass to the subdistribution scale, for two reasons.

i. *Overlapping weighted risk sets*. In the Fine–Gray construction a subject who fails from the competing cause is retained in the cause-of-interest subdistribution risk set (with decreasing weight), and symmetrically. The same subject therefore contributes to *both* event-type score processes, so the cause-1 and cause-2 subdistribution estimators are correlated, 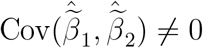, and

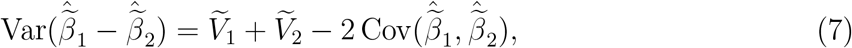

with a covariance term that is not zero and need not be negative.
ii. *Estimated censoring weights*. Both weighted models depend on the *same* estimated *Ĝ*. The naive model-based variance that treats the weights as fixed (as a stacked weighted partial likelihood does by default) omits the variability of *Ĝ* and is generally miscalibrated.

A valid variance for the contrast follows from treating the stacked, weighted estimating equations as an *M*-estimator. Let 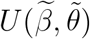 be the stacked weighted score. Because each subject appears in both event-type strata, a sandwich variance that clusters the score contributions on subject captures the cross-stratum covariance in (7). Adding the augmentation term for the estimated weights, the influence function of *Ĝ*, propagated through the weighted score as in the Fine–Gray variance^1,13^, captures source (ii). Writing *ϕ*_*ki*_ for the influence function of 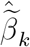 inclusive of the weight-estimation term, the contrast has influence *ϕ*_1*i*_ *− ϕ*_2*i*_ and

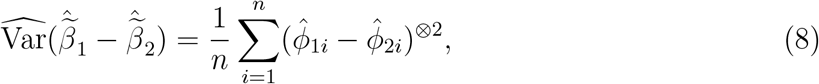

a cluster-robust (on subject), weight-aware sandwich. A nonparametric bootstrap that resamples subjects, re-estimating *Ĝ* within each resample, is an asymptotically equivalent alternative that captures both sources automatically, at higher computational cost. The *ϕ*_*ki*_ are the standard per-cause Fine–Gray influence functions ^1,13^, so the contrast variance (8) follows by differencing them.

The direction of the fixed-weights approximation in source (ii) is known: the influence-function contribution of *Ĝ* enters as a projection of the true-weight score onto the censoring filtration, so estimating the weights reduces the asymptotic variance relative to using the true weights^1^. A subject-clustered sandwich that treats *Ĝ* as fixed therefore captures source (i) exactly and errs on the conservative side for source (ii). Ignoring source (i) by summing the two per-cause variances as if the fits were independent drops the cross-cause covariance, and is anticonservative, as Section 4 confirms.

### 3.3 Stacked pseudo-observation estimation of the cumulative-incidence ratio

Fix a landmark *τ* at which both CIFs are appreciable. Let 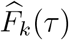 be the Aalen–Johansen estimator^14^ and 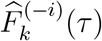 its value with subject *i* deleted. For each cause *k* = 1, 2, subject *i*’s jackknife pseudo-observation is

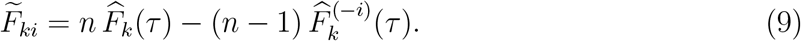

Each 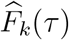 estimates a mean, *E{***1**(*T ≤ τ*, Δ = *k*)*}*, and for such mean-type functionals the pseudo-observations are conditionally unbiased under independent (or covariate-adjusted) censoring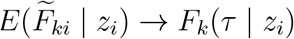^15–17^.

Covariate effects on *ρ*(*τ*) are estimated from a single stacked regression of the 2*n* pseudo-observations. Stack the pairs 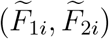 with an event-type indicator *d ∈ {*0, 1*}* (*d* = 1 for the event of interest) and specify the log-link working model

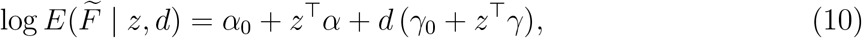

so that the main effects model the nuisance log *F*_2_(*τ* | *z*) and the event-type interactions estimate *γ*_0_ + *z*^*⊤*^*γ* = log *ρ*(*τ* | *z*) directly: *γ* is the covariate effect on log *ρ*(*τ*), and a multi-variate Wald test of *γ* = 0 is the omnibus GCE test on this scale. The estimating function of (10) is linear in the mean-type pseudo-observations, so its conditional unbiasedness (and hence consistency under the working model) follows from the results above. Because each subject contributes two stacked rows, the variance is cluster-robust on subject (Section 3.4). The premise, as for the GCE framework generally, is a genuine competing-risks problem in which both events carry appreciable risk. Where one event is effectively absent, *F*_2_(*τ*) *→* 0 leaves log *ρ* weakly identified: the fit degenerates or returns very large standard errors, which should be read as a signal that the competing-risks premise does not hold.

### 3.4 Variance, and a common loss of orthogonality

Under the conditions of Overgaard et al. ^17^ the per-cause pseudo-observations expand as 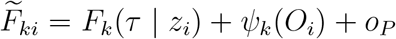 (1), where *ψ*_*k*_ is the influence function of 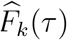; by the delta method, the implied influence function for log *ρ* is *ψ*_*θ*_ = *ψ*_1_*/F*_1_(*τ*) *− ψ*_2_*/F*_2_(*τ*). The stacked GEE estimator of (10) is asymptotically normal with a sandwich variance *A*^*−*1^*BA*^*−*1^ in which each subject’s two stacked score contributions are summed before the outer product, clustering on subject, so the within-subject dependence of the pair 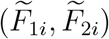 is retained. The precision of a covariate slope on log *ρ* is governed by Var(*ψ*_*θ*_ | *z*), where

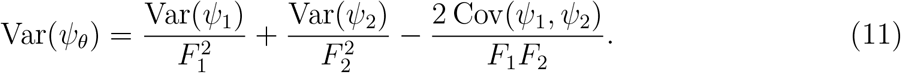

Because the two CIFs are integrals against the same all-cause survivor *S*, their influence functions are negatively associated. A subject who fails of cause 1 at *s ≤ τ* raises *ψ*_1_(*τ*) and, by depleting *S*(*u*^*−*^) in the integrand of *F*_2_ for *u > s*, lowers *ψ*_2_(*τ*). So Cov(*ψ*_1_, *ψ*_2_) *<* 0, the final term of (11) is positive, and the variance of the ratio contrast is inflated relative to the sum of the two single-CIF variances.

For pseudo-observation regressions generally, the usual robust sandwich estimator is known to be biased upward when the working model holds. The jackknife pseudo-observations carry a small negative correlation across subjects that the independence-working sandwich ignores, though the bias is typically modest^18,19^. The subject-clustered variance used here inherits this property: it is expected to err slightly on the conservative side under the model, consistent with the near-nominal calibration observed in Section 4 (SE/SD 0.95–1.05, coverage 0.93–0.95).

The covariance penalty in (11) is the same phenomenon encountered on the subdistribution scale. The cause-specific *ω*^+^ is efficient precisely because its two score processes are orthogonal. Both cumulative-incidence-side contrasts lose that orthogonality and pay a co-variance penalty: 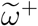 because the weighted subdistribution risk sets overlap across causes and share the estimated censoring weights (7), and *ρ* because the shared survivor couples the two CIFs (11). In each case the covariance term both complicates the variance and bounds the power of the balance test relative to the hazard-scale *ω*^+^. For a divergent effect, the signal in the log-ratio reinforces, of order |*β*_1_| + |*β*_2_|, while the noise grows with the inflated variance. A test on *ρ* (or 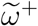) therefore beats the single-event tests when the reinforcement outweighs the inflation, and the advantage shrinks as the two events become more strongly coupled. The interpretability of the cumulative-incidence scales is thus obtained at a quantifiable efficiency cost relative to the cause-specific scale.

## 4 Simulation study

We generate two-group competing-risks data with constant cause-specific hazards *λ*_*k*_(*x*) = *λ*_*k*0_ exp(*β*_*k*_*x*), *x ∈ {*0, 1*}, λ*_10_ = *λ*_20_ = 0.15, and independent exponential censoring with rate 0.15 – equal to the baseline cause hazards, so that approximately one third of subjects in the reference group are censored before either event – evaluating *ρ* and 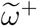 at *τ* = 5. Exponential censoring is used as a simple representative of the independent-censoring condition the estimators require. The applications’ mixture of administrative and dropout censoring satisfies the same condition. Sample sizes were chosen per experiment: *n* = 250 for the null scenario to stress type I control at a moderate sample size, and *n* = 600 for the effect scenarios so that the single-event comparator tests have mid-range power, where comparisons are informative. The null scenario is also reported at *n* = 600 for comparability across rows.

Two questions are examined. First, for the CIF ratio, whether the stacked estimator of Section 3.3 is unbiased with near-nominal coverage and controlled type I error in genuine competing-risks settings, and how its power compares with single-event tests (Table 1).

**Table 1:** Operating characteristics of the stacked cumulative-incidence-ratio estimator (*n*=250 null; *n*=600 divergent, *β*_1_=+0.25, *β*_2_=*−*0.25; 500 replicates). SE/SD is the ratio of the mean cluster-robust standard error to the empirical standard deviation. Single-event power is Gray’s test on each cause.

| Scenario | Estimator | Bias | SE/SD | Coverage | Power<sub>ρ</sub> | Power (single) |
|---|---|---|---|---|---|---|
| Null, *n*=250 | stacked | 0.01 | 0.95 | 0.94 | 0.06 (type I) | 0.06 / 0.04 |
| Null, *n*=600 | stacked | −0.00 | 0.97 | 0.93 | 0.07 (type I) | 0.07 / 0.06 |
| Divergent, *n*=600 | stacked | 0.01 | 0.96 | 0.95 | 0.64 | 0.68 / 0.60 |

Second, for the subdistribution contrast, whether treating the two Fine–Gray models as independent (summing their variances) miscalibrates the standard error of 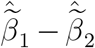, and whether the subject bootstrap, which captures the cross-cause covariance induced by the overlapping subdistribution risk sets, restores nominal coverage (Table 2). This is the empirical test of the variance analysis of Section 3.2. A scope-boundary experiment (shrinking the competing event toward absence) delineates where each estimand is meaningful.

**Table 2:** Calibration of standard errors for the subdistribution 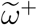 contrast 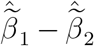 (*n*=600, ~one-third censored; divergent: *β*_1_=+0.5, *β*_2_=*−*0.5; 400 replicates, bootstrap NB=40 datasets *× B*=150 resamples; empirical SD 0.28 in both scenarios). SE/SD is the mean standard error over the empirical standard deviation of the contrast; a value below 1 is anticonservative. The Fine–Gray sum 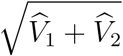 uses each cause’s weight-aware Fine–Gray variance but treats the two fits as independent, dropping the cross-cause covariance. The subject bootstrap captures that covariance (and the estimated weights) and is the reference.

| Scenario | Standard error | Mean SE | SE/SD | Coverage |
|---|---|---|---|---|
| Null | Fine–Gray sum (independent) | 0.198 | 0.71 | 0.86 |
|  | Bootstrap (reference) | 0.263 | 0.94 | 0.95 |
| Divergent | Fine–Gray sum (independent) | 0.210 | 0.76 | 0.83 |
|  | Bootstrap (reference) | 0.269 | 0.97 | 0.93 |

Table 1 confirms approximate unbiasedness (|bias| *≤* 0.01), calibrated standard errors (SE/SD 0.95–0.97), near-nominal coverage (0.93–0.95), type I error consistent with the nominal 0.05 up to Monte Carlo error (0.06–0.07; Monte Carlo SE *≈* 0.01 at 500 replicates), and power comparable to the single-event tests (0.64 versus 0.68/0.60), consistent with the bounded, negative-covariance efficiency trade-off of Section 3.4. The null-scenario rows at *n* = 250 and *n* = 600 are essentially identical, confirming that calibration is not sample-size dependent over this range. In additional runs with large divergent effects (*β*_1_=+1, *β*_2_=*−*1, *n*=600, so the true contrast is *θ*_0_ = 2.0) the stacked estimator remained essentially unbiased (bias 0.01) with calibrated standard errors (SE/SD 1.05) and near-nominal coverage (0.95), confirming that its validity does not degrade with effect size.

Table 2 shows the subdistribution variance analysis of Section 3.2. Summing the two Fine–Gray variances is markedly anticonservative (coverage 0.83–0.86 and SE/SD *≈* 0.71– 0.76) because it omits the cross-cause covariance induced by the overlapping subdistribution risk sets. The subject bootstrap, which captures that covariance (and the estimated weights), restores near-nominal coverage (0.93–0.95, SE/SD *≈* 0.95). Because the empirical standard deviation (0.28) exceeds the independence sum (*≈* 0.20), the covariance is negative and substantial, roughly doubling the contrast variance. This is the subdistribution counterpart of the shared-survivor inflation on the *ρ* scale (Section 3.4), and a caution against reporting a stacked 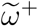 contrast with an independence-based variance.

### Scope of the cumulative-incidence ratio

The premise of these estimators is a genuine competing risk, in which both events carry appreciable incidence. To delineate the boundary we shrank the competing event of the null model toward absence and recorded the fraction of samples in which the *ρ* fit degenerates (non-convergence, or an effectively unbounded estimate or standard error). Table 3 shows that with a genuine competing risk (here *≈* 180 competing events by *τ*) no samples degenerate and the median standard error is small. As the competing event is made nearly absent, degeneracy rises, exceeding half the samples when on average a single competing event is observed. (Event counts here are *observed* counts by *τ*, i.e. after censoring; when the expected count is one, a majority of samples contain no observed competing event, and the degeneracy fraction reflects that.) This behavior is expected. In practice, in the absence of competing events one would not attempt a ratio analysis in the first place. We report the boundary not as a performance result but as an explicit statement of the method’s domain: *ρ* = *F*_1_*/F*_2_ is not a meaningful estimand when there is no competing event in the denominator, and degeneracy of the fit should be read as that signal.

**Table 3:** Scope boundary for the cumulative-incidence ratio (stacked estimator, null effect, *τ* = 5; 500 replicates): as the competing event vanishes, the fit degenerates.

| Setting | *n* | ≈ competing events by *τ* | % degenerate | median SE |
|---|---|---|---|---|
| Genuine competing risk | 600 | 178 | 0.0 | 0.22 |
| Rare competing event | 200 | 5 | 7.4 | 0.96 |
| Competing risk near-absent | 150 | 2 | 36.8 | 1.43 |
| Competing risk near-absent | 120 | 1 | 56.6 | 1.47 |

## 5 Application

We illustrate on a hypothetical `prostate` cohort bundled with `gcemod` (*n* = 1000), with the event of interest metastasis or prostate-cancer death vs. the competing event non-cancer death, at *τ* = 10 years. Here 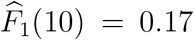 and 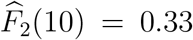, so 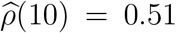: the competing event dominates the realized event mix. Table 5 reports covariate effects on *ρ*(10) from `gcecif()` beside the cause-specific relative *ω*^+^ (`gcecox()`) and the subdistribution 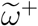 (stacked `gcefg()`). The three contrasts agree in direction but differ in magnitude. For example, Gleason 4+3 gives *ρ* = 2.82, 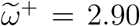, *ω* = 2.15, and comorbidity *ρ* = 0.61, 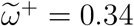, *ω*^+^ = 0.44, reflecting that the cumulative-incidence measures (*ρ* and 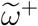) and the cause-specific *ω*^+^ are distinct, non-collapsible summaries of the same balance (Section 2.3).

The cumulative-incidence-side measures are generally more pronounced, e.g. stage T2b+ (*ρ* = 2.29 versus *ω*^+^ = 1.64), though not uniformly: for comorbidity the *ρ* effect (0.61) sits closer to the null than either hazard-scale contrast. Because the stacked working model is log-linear without interactions, the standardized (g-computed) marginal *ρ*(10) ratio of Section 2.5 coincides with the conditional ratio exactly. The multiplicative *F*_2_ weights cancel in the standardization, so the collapsible companion here reproduces the conditional column (comorbidity 0.61; stage 2.29).

A ratio contrast, by construction, cannot distinguish a covariate that affects neither event from one that affects both events equally. For a fuller picture, Table 4 reports the companion reference-event model from the same stacked fit: the main-effects coefficients, which model *F*_2_(10 | *z*), the cumulative incidence of non-cancer death. Age, for example, raises the competing-event incidence by about 3% per year (1.03, 1.02–1.05), while its ratio effect is a modest 0.95. Comorbidity more than doubles the competing-event incidence (2.25, 1.70–2.99), the engine behind its ratio effect of 0.61. The two tables together separate “acts on the balance” from “acts on the rates.”

**Table 4:**
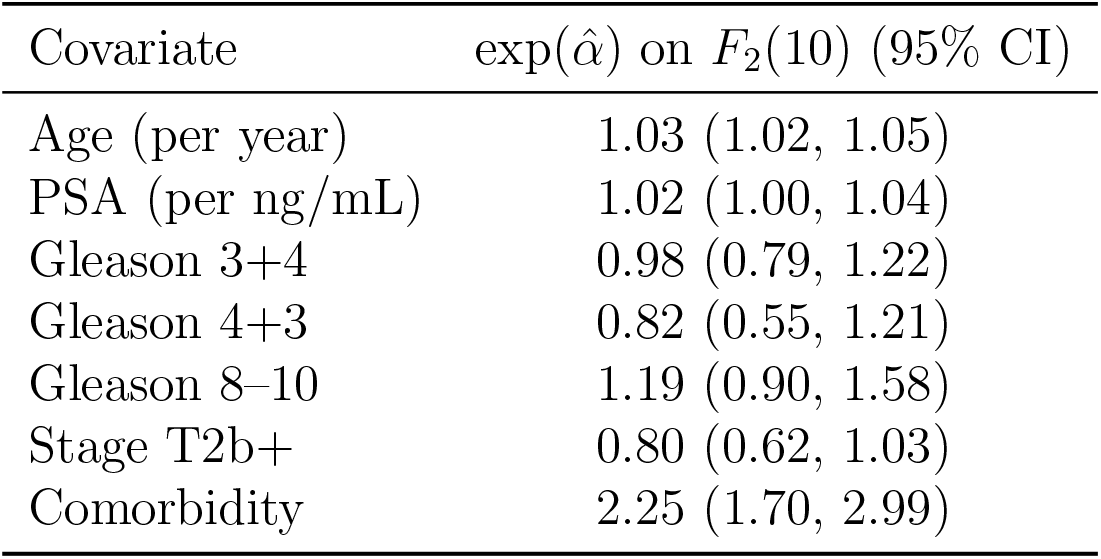
Prostate cohort (*τ* = 10): the companion reference-event model from the same stacked gcecif() fit – multiplicative effects of each covariate on *F*_2_(10 | *z*), the cumulative incidence of the competing event (non-cancer death), from the main-effects block of (10).

| Covariate | exp(α̂) on *F*<sub>2</sub>(10) (95% CI) |
|---|---|
| Age (per year) | 1.03 (1.02, 1.05) |
| PSA (per ng/mL) | 1.02 (1.00, 1.04) |
| Gleason 3+4 | 0.98 (0.79, 1.22) |
| Gleason 4+3 | 0.82 (0.55, 1.21) |
| Gleason 8–10 | 1.19 (0.90, 1.58) |
| Stage T2b+ | 0.80 (0.62, 1.03) |
| Comorbidity | 2.25 (1.70, 2.99) |

**Table 5:**
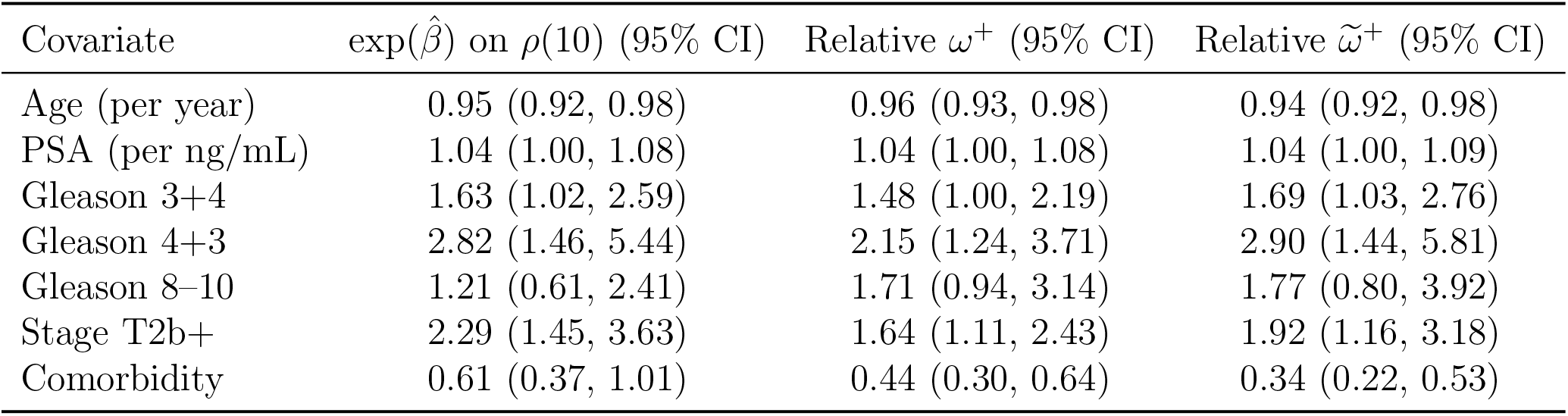
Prostate cohort (*τ* = 10): multiplicative conditional effects on *ρ*(10) (odds the realized event is prostate-cancer death) from the stacked gcecif() pseudo-observation regression, with the cause-specific relative *ω*^+^ for comparison, and the subdistribution 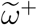 from the stacked gcefg() estimator.

| Covariate | exp(β̂) on ρ(10) (95% CI) | Relative ω<sup>+</sup> (95% CI) | Relative ω̃<sup>+</sup> (95% CI) |
|---|---|---|---|
| Age (per year) | 0.95 (0.92, 0.98) | 0.96 (0.93, 0.98) | 0.94 (0.92, 0.98) |
| PSA (per ng/mL) | 1.04 (1.00, 1.08) | 1.04 (1.00, 1.08) | 1.04 (1.00, 1.09) |
| Gleason 3+4 | 1.63 (1.02, 2.59) | 1.48 (1.00, 2.19) | 1.69 (1.03, 2.76) |
| Gleason 4+3 | 2.82 (1.46, 5.44) | 2.15 (1.24, 3.71) | 2.90 (1.44, 5.81) |
| Gleason 8–10 | 1.21 (0.61, 2.41) | 1.71 (0.94, 3.14) | 1.77 (0.80, 3.92) |
| Stage T2b+ | 2.29 (1.45, 3.63) | 1.64 (1.11, 2.43) | 1.92 (1.16, 3.18) |
| Comorbidity | 0.61 (0.37, 1.01) | 0.44 (0.30, 0.64) | 0.34 (0.22, 0.53) |

Figure 1 shows the `gce omegatime()` diagnostic of Section 6 for this cohort: the non-parametric *ω*^+^(*t*) within GCE risk groups. The group contrast is visibly non-constant over early follow-up. Early events among high-risk patients are predominantly cancer events, with competing mortality accruing only later, and the curves become approximately parallel from mid follow-up onward, with the landmark *τ* = 10 lying within that stable range. The fitted time-constant contrasts are accordingly best read as summaries of the later follow-up, and *ρ*(10) as an average over the realized event mix up to the landmark.

**Figure 1:**
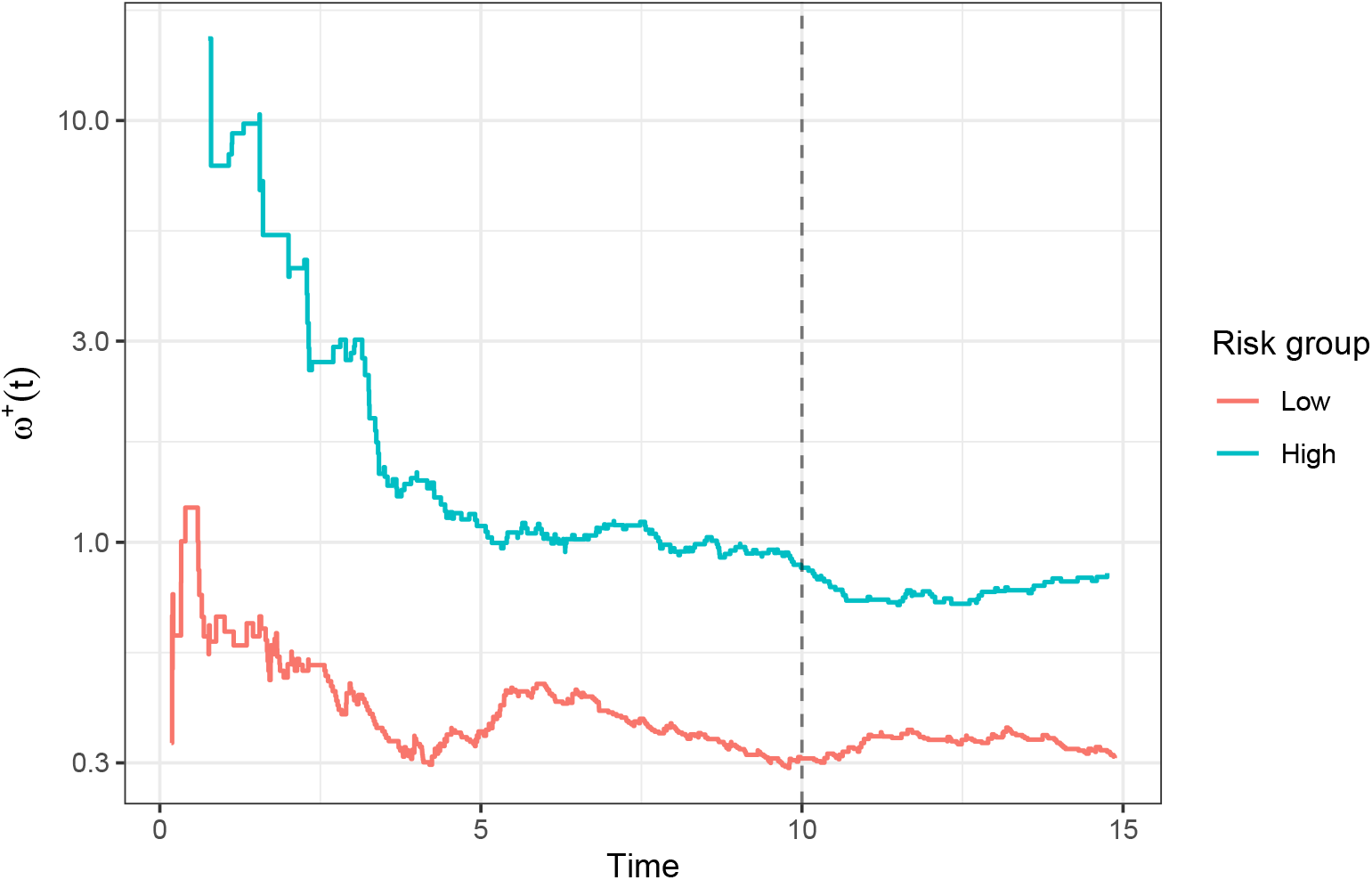
Prostate cohort: Nelson–Aalen estimate of *ω*^+^(*t*) by GCE risk group (log scale), truncated at 15 years; the dashed line marks the landmark *τ* = 10. The group contrast is non-constant over early follow-up and approximately constant thereafter, delineating the range over which the time-constant summaries apply.

A second illustration on a hypothetical head-and-neck cohort (`hn`, *n* = 1000) gives a contrasting scenario: recurrence is the event of interest and the more frequent event, so at *τ* = 5 years 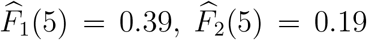, and 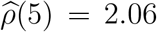– recurrence-dominant, the opposite of the prostate cancer cohort. Table 6 gives the three-scale effects for a parsimonious model (age, smoking, T2–T4 vs. T1, node-positive vs. N0, p16, and oral-cavity site). As in the prostate example, the effects agree broadly in direction but differ in magnitude across the scales. Here, in the higher-incidence cohort, several cumulative-incidence-ratio effects sit closer to the null than their cause-specific counterparts (e.g. p16, *ρ* = 0.76 vs. *ω*^+^ = 0.65; smoker, 0.85 vs. 0.69), consistent with *ρ*(*t*) drifting toward 1 as incidence accumulates (Proposition 2) – the reverse of the prostate cancer cohort, where the cumulative-incidence effects were more pronounced. This cohort is included to illustrate the estimators across contrasting regimes, i.e. recurrence-dominant versus competing event-dominant.

**Table 6:**
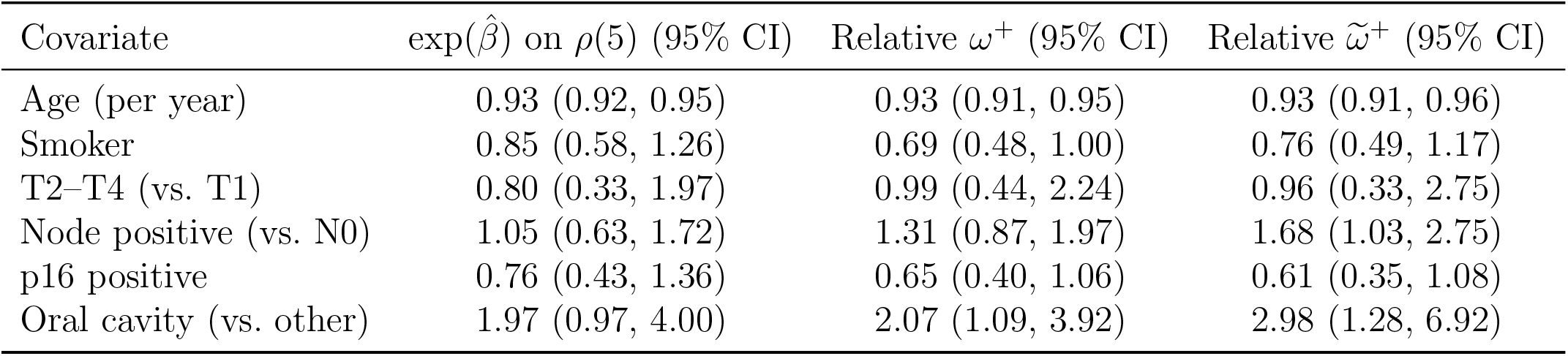
Head-and-neck cohort (*τ* = 5; *n* = 1000, recurrence-dominant, 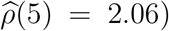: multiplicative effects on *ρ*(5) from the stacked gcecif() estimator, with the cause-specific *ω*^+^ (gcecox()) and subdistribution 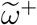 (gcefg()) for comparison, under a parsimonious covariate model.

| Covariate | exp(β̂) on ρ(5) (95% CI) | Relative ω<sup>+</sup> (95% CI) | Relative ω̃<sup>+</sup> (95% CI) |
|---|---|---|---|
| Age (per year) | 0.93 (0.92, 0.95) | 0.93 (0.91, 0.95) | 0.93 (0.91, 0.96) |
| Smoker | 0.85 (0.58, 1.26) | 0.69 (0.48, 1.00) | 0.76 (0.49, 1.17) |
| T2–T4 (vs. T1) | 0.80 (0.33, 1.97) | 0.99 (0.44, 2.24) | 0.96 (0.33, 2.75) |
| Node positive (vs. N0) | 1.05 (0.63, 1.72) | 1.31 (0.87, 1.97) | 1.68 (1.03, 2.75) |
| p16 positive | 0.76 (0.43, 1.36) | 0.65 (0.40, 1.06) | 0.61 (0.35, 1.08) |
| Oral cavity (vs. other) | 1.97 (0.97, 4.00) | 2.07 (1.09, 3.92) | 2.98 (1.28, 6.92) |

## 6 Software

Both estimators are implemented in the `gcemod` R package (version 0.4.0). The stacked subdistribution estimator of 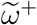 is provided by `gcefg()` (with the cause-specific *ω*^+^ analogue `gcecox()`), and the stacked pseudo-observation estimator of the cumulative-incidence ratio *ρ* by `gcecif()`. Each returns a common `gcemod` object with the coefficient table on its scale, an omnibus test, and shared risk-score and stratification tooling. On the subdistribution scale, the default standard errors are the subject-clustered sandwich with the censoring weights treated as fixed, and the subject bootstrap is available via `se = “boot”`. The subject-clustered pseudo-observation variance of Section 3.4 is the standard error reported by `gcecif()`. A diagnostic display, `gce omegatime()`, plots a nonparametric estimate of a user-selected GCE contrast – *ω*(*t*), *ω*^+^(*t*), their subdistribution analogues, *ρ*(*t*), or its bounded share form *π*(*t*) = *ρ*(*t*)*/{*1 + *ρ*(*t*)*}* – over follow-up, overall or within risk groups, as a visual assessment of the proportional relative-hazards assumption and of the time range over which it holds approximately.

## 7 Discussion

We have applied the GCE contrast from the cause-specific hazard scale to the two scales that describe realized incidence. We developed a stacked, risk-set-weighted Lunn–McNeil estimator for the subdistribution hazard ratio 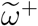 and a stacked pseudo-observation estimator for the cumulative-incidence ratio *ρ*, so that on all three scales the contrast is read off a single stacked fit: an event-type indicator, nuisance main effects, and interaction coefficients that are the contrast, with a subject-level variance. Relating the three (Section 2.3) shows they are distinct summaries of one balance – *ρ* equal to *ω*^+^ exactly under proportional cause-specific hazards, and to 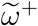 only in the small-time limit under proportional subdistribution hazards – so reporting more than one is informative. The drift of *ρ* toward unity as incidence accumulates (Proposition 2) shows why the net effect of treatment on a composite endpoint attenuates toward no effect under heavy competing risks, as observed previously^10^. The unifying methodological point is that the orthogonality that makes the cause-specific contrast efficient is lost on both cumulative-incidence scales: 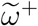 couples the two causes through over-lapping weighted risk sets and shared censoring weights, and *ρ* through the shared survivor. In each case the resulting covariance dictates a different handling of the variance – cluster-robust and weight-aware for 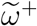, subject-clustered across the per-cause pseudo-observations for *ρ* – and bounds the efficiency of the balance test relative to the hazard scale.

Two prior cumulative-incidence quantities are related but distinct: the Pepe–Mori conditional probability *F*_1_*/*(1 *− F*_2_)^8^ conditions on *not* having had the competing event, whereas *π*_1_ conditions on having had *some* event. Vertical modeling^9^ targets the *instantaneous* relative cause probability, whereas *ρ*(*t*) is *cumulative*. Which scale to prefer depends on the goal: the cause-specific *ω*^+^ for maximal power to detect a balance shift, the subdistribution 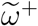 and cumulative-incidence *ρ* for statements aligned with realized incidence, at the cost of efficiency.

The interpretability of subdistribution-hazard regression itself has been debated. The subdistribution risk set retains subjects who have already failed from the competing event, so its coefficients do not describe event rates in any observable surviving population^20^. However, a principal goal of GCE modeling is the construction of classifiers that identify subgroups that are more (or less) likely to benefit from treatment intensification, validated empirically by testing the interaction of the treatment effect with the proposed classifier. Such a classifier could be built on any of the three scales, and which scale yields the best-performing classifier is an empirical question for the application at hand. It would be difficult to establish that the cause-specific scale categorically outperforms the subdistribution scale for prediction in all cases. The subdistribution hazard scale may be preferred when realized incidence is clinically the most relevant. Providing a valid methodology on all three scales also allows for such empirical comparison.

Note that the GCE contrast carries signal only when the two events respond differently to the covariates, i.e. the events have distinct etiologies with contrasting risk-factor profiles, as often arises, for example, for cancer-specific events versus non-cancer mortality in oncology. When the competing events are mechanistically similar, the contrast is null by construction, *ω*^+^ varies little across patients, and the risk score is flat. Thus, *ω* guides selection when *θ*_1_*≠ θ*_2_, and the same distinctness of cause-specific effects is what makes covariate effects on the balance nonzero. Model degeneracy is thus not a failure but diagnostic. Uniformly null GCE contrasts indicate that the two events are not distinct enough to warrant separate modeling, and a pooled analysis loses nothing. Importantly, the condition requires no assumption about dependence between the latent event times.

Model checking for the proposed estimators has three natural components. First, the time-constancy that makes a single summary adequate can be assessed visually with the nonparametric contrast-over-time displays of Section 6 (Figure 1): a flat *ω*^+^(*t*) or *ρ*(*t*) supports the proportional relative-hazards premise, and parallel group curves on the log scale support a time-constant covariate contrast even where the individual curves drift. Second, the landmark *τ* is an analysis choice; refitting (10) over a grid of landmarks shows whether conclusions are sensitive to it (under proportional relative hazards, *ρ*(*τ* | *z*) ratios are constant in *τ*, so stability across landmarks is itself a check of the premise). Third, the log link in (10) is a working-model choice on the multiplicative scale; standard pseudo-observation residual diagnostics^15^ apply to the stacked fit, and gross link misspecification manifests as systematic trends in the pseudo-observation residuals against the linear predictor.

Finally, the construction generalizes beyond two competing events. With *K* causes, the stacked regression accommodates *K* per-cause pseudo-observations per subject with cause indicators, and the interaction blocks estimate log*{F*_*k*_(*τ* | *z*)*/F*_*K*_(*τ* | *z*)*}* against a chosen reference cause – a multinomial analogue in which any ratio of interest follows by contrast. In applications with several events of interest, the practically useful contrasts are likely to be composite-versus-composite, i.e. the pooled events of interest against the pooled competing events, as in the GCE tradition, or each cause against the shared competing event. The two-event case developed here is the one with the clearest clinical interpretation, and we leave the multi-cause extension to future work.

## Data Availability

All data are available via the gcemod package.

## Data and code availability

The hypothetical example cohorts (which emulate clinical data but are not from actual subjects) and both estimators (`gcefg()`, `gcecif()`) are distributed in the `gcemod` R package. A replication script reproducing all results is provided as supplementary material.

